# Timeliness of reporting of SARS-CoV-2 seroprevalence results and their utility for infectious disease surveillance

**DOI:** 10.1101/2022.02.17.22271099

**Authors:** Claire Donnici, Natasha Ilincic, Christian Cao, Caseng Zhang, Gabriel Deveaux, David A. Clifton, David Buckeridge, Niklas Bobrovitz, Rahul K. Arora

**Affiliations:** Cumming School of Medicine, University of Calgary, Calgary, AB, Canada, 3330 Hospital Dr NW, Calgary, AB T2N 4N1; Temerty Faculty of Medicine, University of Toronto, Toronto, ON, Canada, 27 King’s College Cir, Toronto, ON M5S 1A1; Faculty of Health Sciences, McMaster University, Hamilton, ON, Canada, 1280 Main St W, Hamilton, ON L8S 4L8; Institute of Biomedical Engineering, University of Oxford, UK, Old Road Campus Research Building, Headington, Oxford OX3 7DQ; Faculty of Medicine and Health Sciences, McGill University, Montreal, QC, Canada, 3605 Rue de la Montagne, Montreal, QC H3G 2M1

**Author notes:** **Corresponding author:** Claire Donnici, MD/MSc Student, University of Calgary, Calgary, AB, Canada, SeroTracker Research Team. Claire Donnici and Natasha Ilincic are co-first authors on this manuscript.

**Keywords:** Public health surveillance, seroprevalence, infectious disease, COVID-19, reporting, bibliometrics

## Abstract

Seroprevalence studies have been used throughout the COVID-19 pandemic to monitor infection and immunity. These studies are often reported in peer-reviewed journals, but the academic writing and publishing process can delay reporting and thereby public health action. Seroprevalence estimates have been reported faster in preprints and media, but with concerns about data quality. We aimed to (i) describe the timeliness of SARS-CoV-2 serosurveillance reporting by publication venue and study characteristics and (ii) identify relationships between timeliness, data validity, and representativeness to guide recommendations for serosurveillance efforts.

We included seroprevalence studies published between January 1, 2020 and December 31, 2021 from the ongoing SeroTracker living systematic review. For each study, we calculated timeliness as the time elapsed between the end of sampling and the first public report. We evaluated data validity based on serological test performance and correction for sampling error, and representativeness based on use of a representative sample frame and adequate sample coverages. We examined how timeliness varied with study characteristics, representativeness, and data validity using univariate and multivariate Cox regression.

We analyzed 1,844 studies. Median time to publication was 154 days (IQR 64-255), varying by publication venue (journal articles: 212 days, preprints: 101 days, institutional reports: 18 days, and media: 12 days). Multivariate analysis confirmed the relationship between timeliness and publication venue and showed that general population studies were published faster than special population or health care worker studies; there was no relationship between timeliness and study geographic scope, geographic region, representativeness, or serological test performance.

Seroprevalence studies in peer-reviewed articles and preprints are published slowly, highlighting the limitations of using the academic literature to report seroprevalence during a health crisis. More timely reporting of seroprevalence estimates can improve their usefulness for surveillance, enabling more effective responses during health emergencies.

## Introduction

Timely information about population immunity can be critical for effective public health decision making, as emphasized throughout the COVID-19 pandemic. Seroprevalence studies estimate the prevalence of antibodies and are crucial sources of this information. Estimates of seroprevalence can inform scenario modeling, public health planning, and national policies.

During the COVID-19 pandemic, seroprevalence estimates have primarily been generated through standalone research studies instead of ongoing public health surveillance efforts, raising questions about their public health impact.^1–3^ For these seroprevalence studies to be effective when used for public health surveillance, it is important that they have the attributes of effective surveillance systems — including timeliness, representativeness, and validity, among others (Table 1).^4,5^ However, many of these attributes are challenging to realize through one-off study efforts, particularly when findings are shared in research manuscripts published as peer-reviewed articles.

**Table 1.** Attributes of effective public health surveillance systems used in this study and the measures used to examine them.

| Attribute<sup>a</sup> | Definition | Measures used in this study<sup>b</sup> |
| --- | --- | --- |
| Timeliness | Refers to the time between any two steps in the surveillance system. For example, the time between the onset of a health event and reporting of the event to a public health agency.<br>Timeliness can be evaluated by the availability of information for control of health-related events and is influenced by surveillance methods and data sources.<sup>4</sup> | **How rapidly were results released after participants were sampled?** The time elapsed between the last date of participant sample collection (sampling end date) and the date of the release of results from a study, irrespective of type of publication platform. For studies released via multiple platforms (e.g., government report, preprint and journal article), we used the first date the results were available. |
| Data quality (validity)<sup>c</sup> | Refers to the validity of data in the surveillance system, which is influenced by the performance of screening tests<sup>5</sup>, statistical methods, and surveillance methods (i.e., study type, geographical scope, etc.).<br>Validity refers to the proportion of data entries that correctly reflect the true value of the data collected.<sup>4</sup> | **Were valid methods used to identify seropositivity?** Did the serology test used meet the FDA standards for Emergency Use Authorization for COVID-19 serology tests, with sensitivity $\geq 90\%$ and specificity $\geq 95\%$?<br>**Was there a correction for the sampling error?** If non-probability sampling was used, then were statistical adjustments or reweighting of sample demographics performed? |
| Representativeness<sup>c</sup> | Refers to the ability of the surveillance system to accurately describe the health event over time.<br>This is achieved by considering its distribution in the population. Surveillance data should be described in terms of geography, demographics, and clinical manifestations.<sup>4</sup> | **Was the sample frame appropriate for the study to generalize its findings to the population of interest?** Was the sample used in the study generally representative of the target population? As an example, a sample of healthcare workers in predominantly administrative roles would not be representative of all healthcare workers working during the pandemic.<br>**Was data analysis conducted with sufficient coverage of the identified sample?** To evaluate coverage, we examined if the demographics of a sample aligned with the expected demographics in the target population (age, sex, ethnicity). |

<sup>a</sup>Other attributes of effective public health surveillance not evaluable based on seroprevalence study reports and not examined in detail here include simplicity, flexibility, acceptability, stability, sensitivity, and positive predictive value.<sup>4,5</sup>
<sup>b</sup>The response options for each measure were “Yes”, “No”, or “Unclear”.
<sup>c</sup>These items were derived from a modified version of the Joanna Briggs Institute Critical Appraisal Checklist for Prevalence Studies.

**Table 2.** Cox Regression Table. Hazard ratios > 1 mean that the comparator group was published faster than the control group. Univariate models were conducted using Cox proportional hazards models with a single predictor of timeliness. Variables that were significant predictors of timeliness in univariate analysis were included in multivariate Cox regression. Overall risk of bias was not included to avoid collinearity with individual items evaluating data representativeness and quality. The overall multivariate p value was <2e-16 according to the Wald test.

| Reference group | Comparison group | Univariate Cox regression hazard ratio [95% CI] | Univariate Cox regression p value | Multivariate Cox regression hazard ratio [95% CI] | Multivariate Cox regression p value |
| --- | --- | --- | --- | --- | --- |
| **Publication Venue - *Journal article (peer-reviewed)*** | Media | 20.4 [16.4-25.3] | <2e-16*** | 17.0 [13.3-21.8] | <2e-16*** |
|  | Institutional Report | 12.2 [10.1-14.8] | <2e-16*** | 12.4 [10.1-15.1] | <2e-16*** |
|  | Preprint | 2.3 [2.0-2.5] | <2e-16*** | 2.2 [2.0-2.5] | <2e-16*** |
|  | Presentation or Conference | 1.3 [1.0 - 1.8] | 0.05 | 1.3 [0.96 - 1.7] | 0.09 |
| **Sample Frame - *Household and community samples*** | Blood donors or residual sera | 0.76 [0.65 - 0.88] | 3.7e-04*** | 0.88 [0.74 - 1.05] | 0.15 |
|  | Healthcare workers and caregivers | 0.58 [0.51 - 0.66] | <2e-16*** | 0.81 [0.70-0.94] | 0.004** |
|  | Other special populations | 0.58 [0.51 - 0.66] | <2e-16*** | 0.74 [0.64-0.86] | 6e-05*** |
|  | Multiple populations | 0.69 [0.58 - 0.82] | 1.7e-05*** | 0.90 [0.74 - 1.08] | 0.26 |
| **WHO Region - *AMRO*** | AFRO | 1.13 [0.89 - 1.43] | 0.30 | 1.17 [0.92-1.49] | 0.19 |
|  | EMRO | 0.77 [0.62 - 0.95] | 0.02* | 0.84 [0.68-1.05] | 0.14 |
|  | EURO | 0.85 [0.77 - 0.95] | 0.004** | 0.95 [0.85-1.06] | 0.33 |
|  | SEARO | 1.09 [0.91 - 1.31] | 0.35 | 1.03 [0.86-1.25] | 0.73 |
|  | WPRO | 0.87 [0.72 - 1.06] | 0.17 | 0.90 [0.74-1.10] | 0.30 |
| **Was the sample representative of the target population? - *Yes*** | No | 0.96 [0.87-1.06] | 0.43 | 1.10 [0.99 -1.22] | 0.09 |
|  | Unclear | 1.30 [1.11-1.53] | 0.001** | 1.07 [0.90-1.27] | 0.45 |
| **Was there appropriate sample coverage? - *Yes*** | No | 0.83 [0.71-0.98] | 0.03* | 0.85 [0.72-1.00] | 0.06 |
|  | Unclear | 1.04 [0.93-1.16] | 0.48 | 1.01 [0.90 -1.14] | 0.82 |
| **Did the antibody test used meet FDA EUA standards? - Yes** | No | 1.09 [0.91-1.32] | 0.35 | 1.01 [0.83-1.22] | 0.92 |
|  | Unclear | 1.24 [1.13-1.36] | 8.5e-06*** | 1.08 [0.98-1.19] | 0.14 |
| **Was appropriate sampling or a population adjustment performed? -Yes** | No | 0.77 [0.67-0.89] | 3.6e-04*** | 1.20 [1.03-1.41] | 0.02* |
| **Geographical Scope - National** | Regional | 0.88 [0.75 - 1.03] | 0.099 | Not included | Not included |
|  | Local | 0.93 [0.82 - 1.05] | 0.23 | Not included | Not included |
| **Overall Risk of Bias - Low** | Moderate | 1.13 [0.87 - 1.47] | 0.34 | Not included | Not included |
|  | High | 1.12 [0.87 - 1.44] | 0.38 | Not included | Not included |
|  | Unclear | 2.22 [1.65 - 2.99] | 1.2e-07*** | Not included | Not included |
| **Levels of Significance \* - < 0.05; \*\* < 0.01; \*\*\* - < 0.001** |  |  |  |  |  |

Peer-review can delay the availability of data by months. The scientific publication process has been criticized for these delays,^6–8^ as these have an impact on public health responses, and can also hinder secondary analysis, modeling, and global comparisons. At the same time, journals are not designed for the routine reporting of surveillance data and may not even consider updated results as sufficiently novel for publication.

To expedite dissemination of results, some researchers have turned to more rapid and accessible platforms, such as news and media,^9^ government reports,^10^ and preprints.^11^ However, the generalizability and validity of such non-peer-reviewed evidence has been questioned.^12,13^ It remains unclear whether these alternative platforms do indeed lead to faster reporting compared to scientific journals, and whether the ability to bypass peer-review has resulted in prolific publication of weaker evidence.

We aimed to determine the timeliness of SARS-CoV-2 seroprevalence studies in providing information useful for public health surveillance and further analysis. To do so, we analyzed a global database of SARS-CoV-2 seroprevalence studies, aiming to:

1. describe the timeliness of SARS-CoV-2 seroprevalence reporting by publication venue, study methods, and populations studied
2. identify whether more timely reporting compromises other facets of effective surveillance, by examining relationships between timeliness, data quality, and representativeness.

## Methods

### Study identification, data extraction, and quality assessment

We identified SARS-CoV-2 seroprevalence studies using a living systematic review registered with PROSPERO (CRD42020183634).^1^ Data sources and searching methods have been previously described.^2^ In brief, we conducted a search of electronic databases, grey literature, and news media for cohort and cross-sectional studies reporting seroprevalence estimates published between January 1, 2020-December 31, 2021. We also invited submissions of seroprevalence studies on our dashboard, at SeroTracker.com.^14^

Inclusion criteria, screening, data extraction and quality assessment of seroprevalence studies have also been previously described in detail.^2^ We included SARS-CoV-2 seroprevalence studies in humans. To be included, studies had to report a sample size, sampling end date, geographic location of sampling, and a seroprevalence estimate. All records were screened independently and in duplicate. A risk of bias (RoB) assessment was performed by two independent reviewers. The assessment involved use of a modified nine-item Joanna Briggs Institute (JBI) Critical Appraisal Checklist for Prevalence Studies and, based on the results of the JBI checklist, generation of an overall RoB rating (low, moderate, high, unclear)^15–17^.

For all included studies, we identified the first date at which results were published after data collection ended, irrespective of publication venue. For each study, we categorized publication venue as a peer-reviewed journal article, preprint, institutional report (a report from a government, organization or institution presenting data in a formal but non-academic manuscript format), presentation or conference materials (abstracts, PDF presentations) or media (media releases and news reports). We categorized sample frame as (i) household or community samples, (ii) blood donors or residual sera, (iii) healthcare workers, (iv) other special populations, which included essential non-healthcare workers, non-essential workers, students and daycares, vulnerable individuals (persons who are incarcerated, persons who are experiencing homelessness), non-COVID-19 patients and hospital visitors, and (v) studies that sampled multiple different populations.

### Defining study timeliness, representativeness, and data quality

In this work, we focused on timeliness as a key determinant of the effectiveness of seroprevalence data for public health surveillance. We also examined the relationships between timeliness and two other characteristics of effective public health surveillance: representativeness and data quality. Table 1 provides definitions of each attribute and the measures used to operationalize them in the present study.

### Analysis

We calculated the median publication timeliness of seroprevalence studies with IQR in both the overall sample and stratified by publication venue (peer-reviewed journal articles, preprints, presentation or conference materials, institutional reports and media). We compared the median timeliness between preprints and institutional reports or media, and between peer-reviewed publications and preprints.

To assess the role of preprints in expediting the release of data, we compared the median time to publication for peer-reviewed journal articles that first appeared as preprints and peer-reviewed journal articles that were not preprinted.

We examined the relationship between timeliness and study characteristics (i.e., publication venue, geographic scope, sample frame, WHO region, and overall RoB), each measure of representativeness, and each measure of validity. To do so, we generated stratified Kaplan-Meier plots and conducted univariate Cox regressions, calculating overall model p values with the Wald test. To directly compare timeliness between publication venues, we conducted pairwise log-rank tests, using the Bonferroni correction to adjust for multiple comparisons. The reference groups chosen for Cox regression were as follows: geographic scope - “national”, WHO region - “Region of the Americas (AMRO)”, sample frame - “household and community samples”, overall RoB - “low”, individual items evaluating data representativeness and validity (i.e., was the sample representative of the target population) - “yes”.

To examine which factors were independently associated with timeliness, we constructed a multivariate Cox model. The predictors used in this model were all those which were significant on univariate Cox regression. Overall RoB was excluded from the multivariate model because it is partially determined by the measures of representativeness and data quality we employed. Analysis was completed using the survminer library and ggsurvplot function in R (Version 4.0.5).

## Results

Overall, 1,844 studies were included in the analysis. The majority (59%) of studies were first released as peer-reviewed journal articles, followed by preprints (24.2%), institutional reports (7.81%), news articles (6.24%), and presentations or conference abstracts (2.66%). The majority (78%) of studies were single time point (cross-sectional studies) as opposed to studies with repeated measures.

Across all publication venues, median time to publication was 154 days (IQR: 64 - 255). The shortest time to publication was 0 days for a media report, while the longest was 556 days for a peer-reviewed article.

Timeliness varied significantly across publication venues (Figure 1). Media reports (median: 12 days; IQR: 3-25) were released significantly faster than institutional reports (median: 18 days; IQR: 2 - 45) (log-rank p = 0.02). Both media and institutional reports were published significantly faster than studies released in all other publication venues (log-rank p < 2e-16). Preprints (median: 101 days; IQR: 49 - 180) were released faster than presentation or conference materials (median: 187 days; IQR: 43 - 295) (log-rank p = 0.003), and both venues released study results in significantly faster time compared to peer-reviewed journal articles (median: 212 days; IQR: 131 - 305) (log-rank p < 2e-16 & log-rank p = 0.049, respectively).

**Figure 1.**
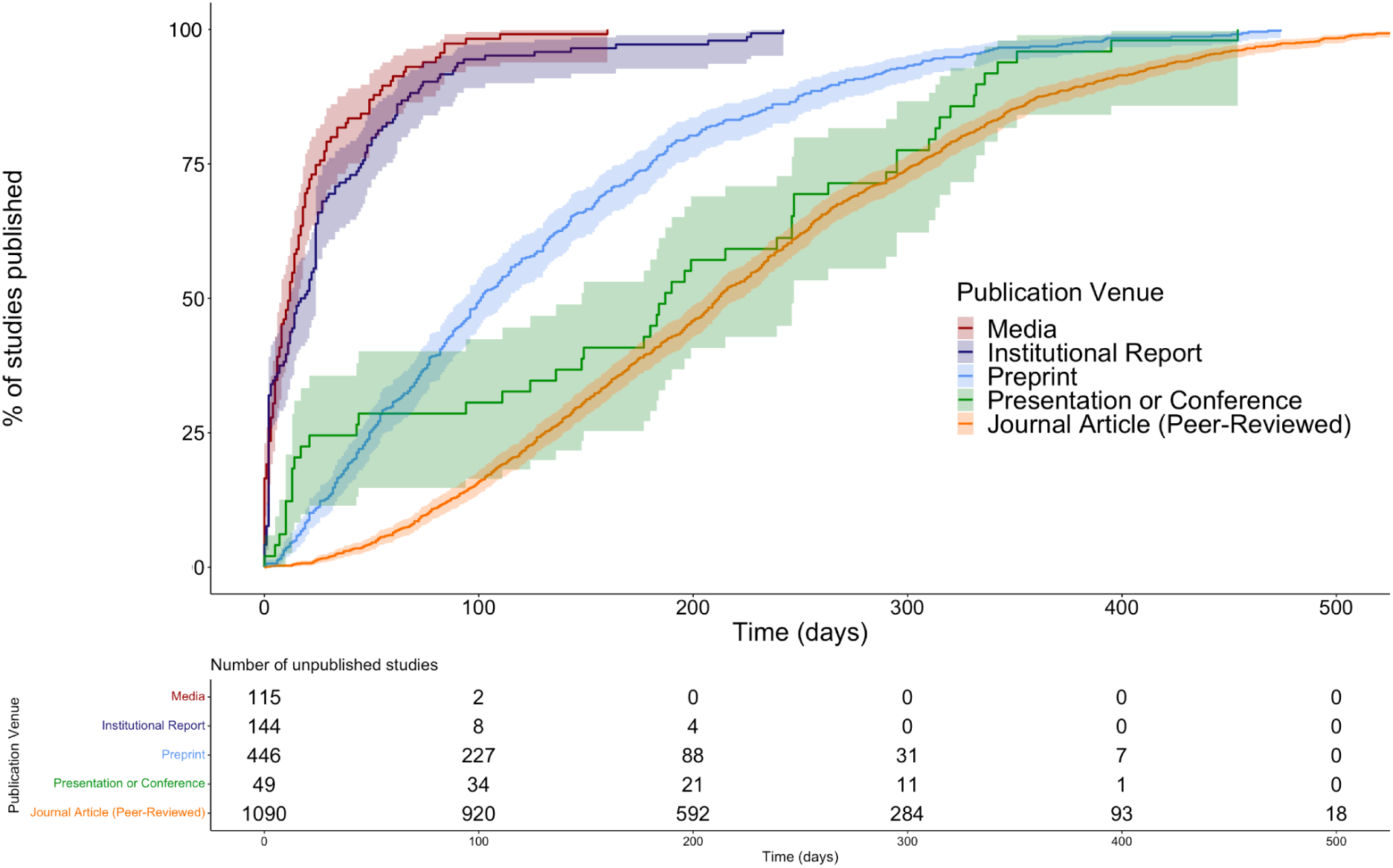
Kaplan-Meier curve and risk table for timeliness by publication venue. Pairwise comparisons indicate significant differences in timeliness between publication venues. Media and institutional reports were published significantly faster than all other publication venues (all log-rank p < 2e-16, with Bonferroni correction). Preprints were published in significantly shorter time compared to journal articles (log-rank p < 2e-16) and presentation or conference materials (log-rank p = 0.003). Presentation and conference materials were also released faster than journal articles (log-rank p = 0.049). Timeliness curves are plotted with 95% confidence intervals (shaded area).

There were 230 studies first published as preprints that later appeared as peer-reviewed journal articles. There was no significant difference in time to publication of these studies and studies that were released as peer-reviewed journal articles without preprinting (p = 0.3).

Examination of RoB by publication venue showed that the fewest low or moderate risk of bias studies appeared in presentation or conference materials (5.1%) and media reports (9.6%). There were larger proportions of low or moderate RoB studies reported in peer-reviewed journal articles (32%), preprints (42%), and institutional reports (51%). Media reports had the highest number of studies that had insufficient information to evaluate bias (40% unclear RoB).

Compared to AMRO, there were significant differences in timeliness for studies conducted in different WHO regions (overall p = 0.002). This result was driven by slower study publication in European Region (EURO) as compared to AMRO (hazard ratio [HR] = 0.85, 95% confidence interval [0.77-0.95], p = 0.004) and the Eastern Mediterranean Region (EMRO) (HR 0.77 [0.62-0.95], p = 0.02) as compared to AMRO (Figure 2A). We observed significant differences in timeliness by overall RoB (overall p = 8 e-14), driven by significantly faster timeliness for studies at unclear RoB (i.e., insufficient information to evaluate) as compared to low RoB (HR 2.22 [1.65-2.99], p = 1.2e-07), whereas there were no differences between studies at moderate (values) or high (values) RoB vs. low RoB (Figure 2B). There were significant differences in timeliness by sample frame (overall p = <2e-16), where studies of blood donors or residual sera (HR 0.76 [0.65 - 0.88], p = 3.7e-04), multiple populations (HR 0.69 [0.58 - 0.82], p = 1.7e-05), healthcare workers (HR 0.58 [0.51 - 0.66], p = <2e-16), and other special populations (HR 0.58 [0.51 - 0.66], p = <2e-16), took significantly longer to be released than studies of household/community samples (Figure 2C). There were no differences in timeliness by geographic scope of a study (overall p = 0.3) (Figure 2D).

**Figure 2.**
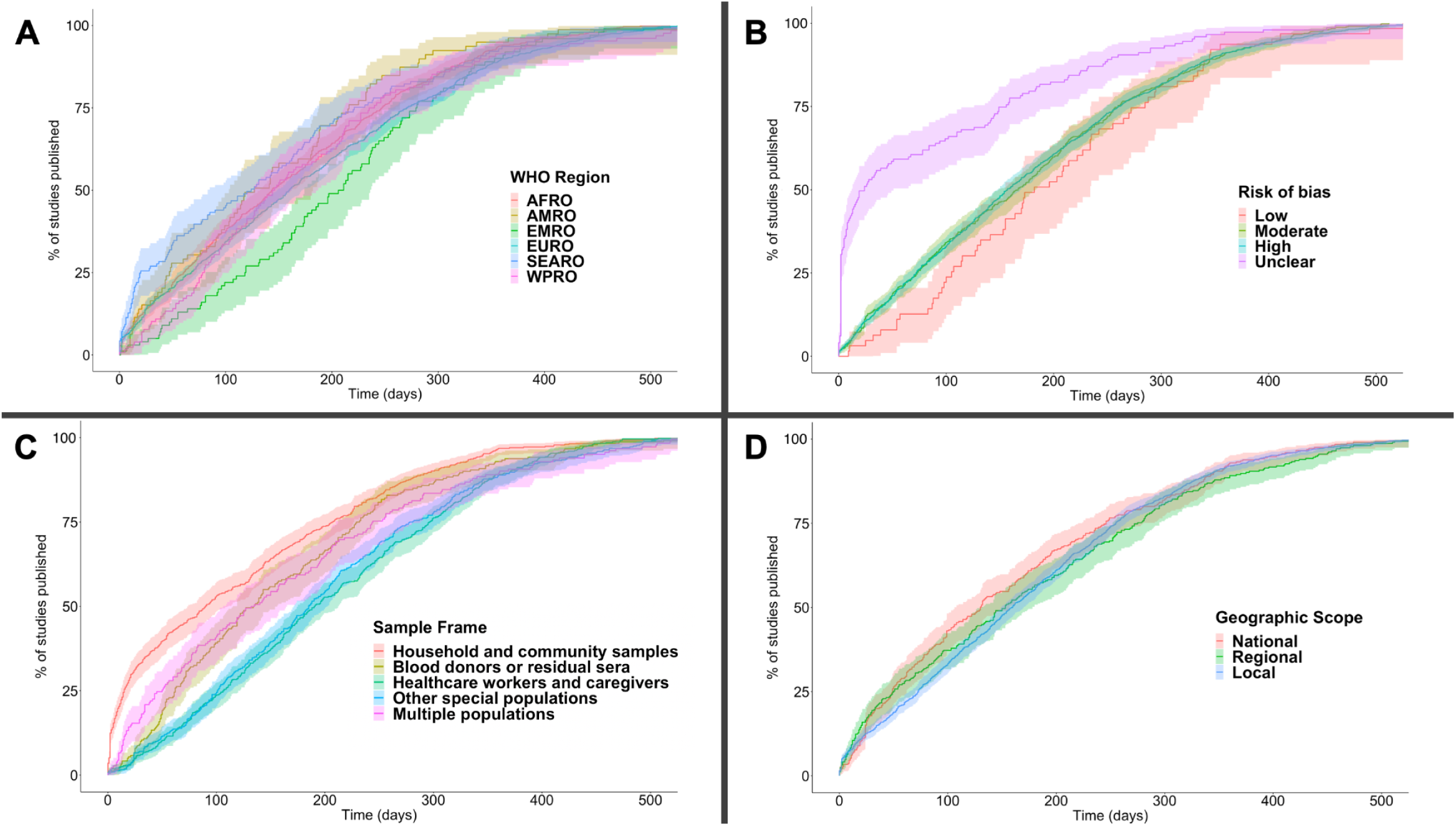
Kaplan-Meier curves for timeliness across study characteristics. Comparison of study timeliness according to (A) the WHO region the study was conducted in (reference: AMRO; overall p = 0.002), (B) overall risk of bias (reference: low; overall p = 8e-14), (C) sample frames (reference: household and community samples; overall p = <2e-16) and (D) geographic scope (reference: national; overall p = 0.3). Timeliness curves are plotted with 95% confidence intervals (shaded area).

Studies that did not have sufficient information to evaluate if the study sample was representative of the target population (HR 1.30 [1.11 - 1.53], p = 0.001) (Figure 3A) and studies that did not report antibody test sensitivity and specificity (“unclear”) (HR 1.24 [1.13 - 1.36], p = 8.5e-06) (Figure 3C) had a higher probability of publication in shorter time compared to studies that reported these data and met the criteria for representativeness and high test sensitivity and specificity. Studies that were not conducted with significant coverage of the sample (HR = 0.83 [0.71 - 0.98], p = 0.03) (Figure 3B) or that did not use either appropriate sampling methods or a population adjustment (HR = 0.77 [0.67 - 0.89], p = 3.6e-04) (Figure 3D) were published slower compared to studies that met these criteria.

**Figure 3.**
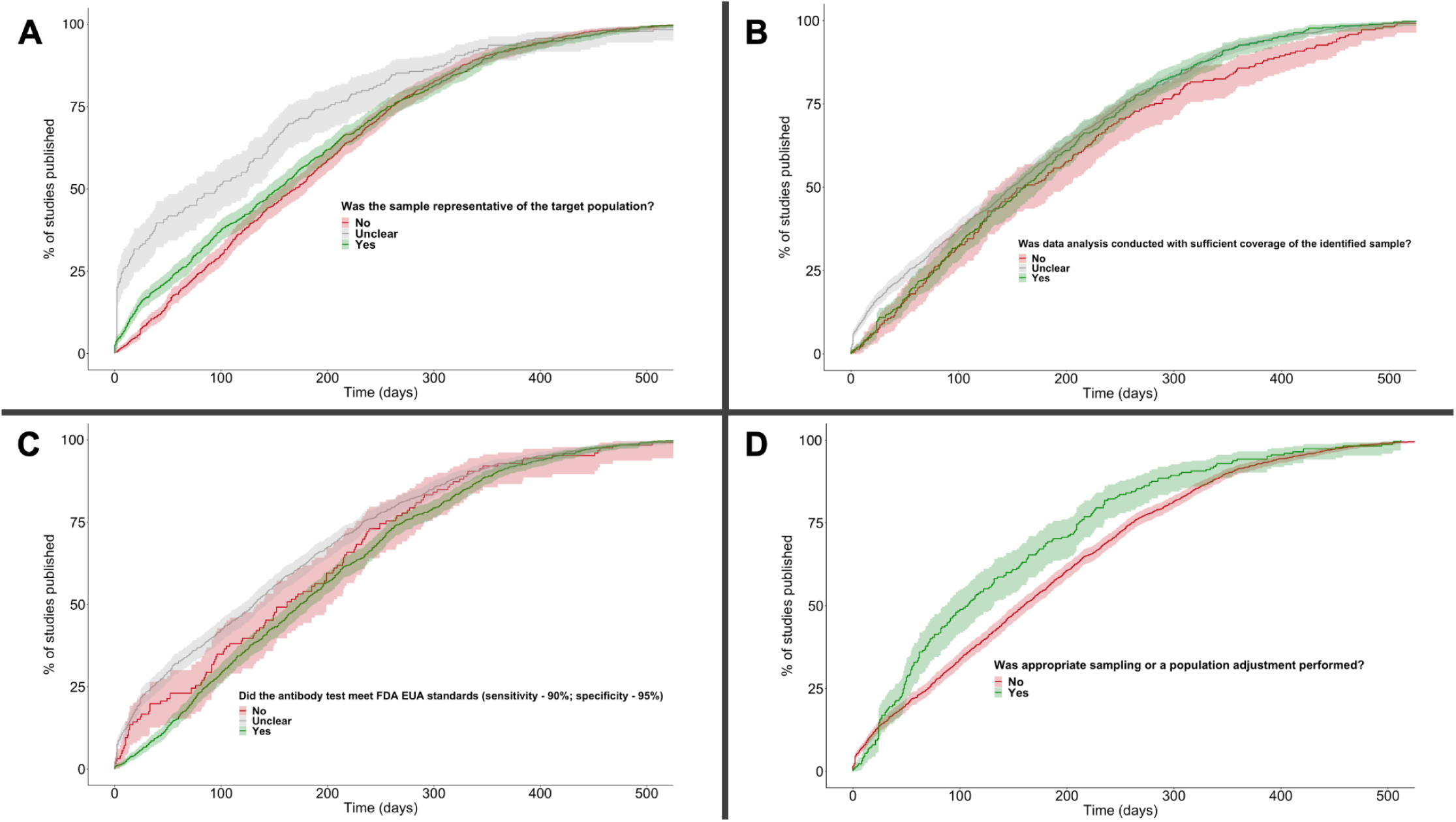
Kaplan-Meier curves for timeliness across measures of study representativeness and data quality. Comparison of timeliness according to (A) whether or not the sample was representative of the target population (overall p = 9e-04), (B) sample coverage (overall p = 0.01), (C) sensitivity and specificity of the antibody test used (overall p = 5e-05) and (D) appropriateness of sampling method and statistical analysis (overall p = 4e-04). The reference group was “yes” for all analyses. Timeliness curves are plotted with 95% confidence intervals (shaded area).

Predictors included in the multivariate Cox regression were all publication venue, sample frame, WHO region, and individual measures of data validity and representativeness. Overall risk of bias was excluded to avoid collinearity with individual measures. Compared to peer-reviewed journal articles, preprints (HR 2.24 [2.0 - 2.52], p = <2e-16), media (HR 17.0 [13.3 - 21.8], p = <2e-16), and institutional reports (HR 12.4 [10.1 - 15.1], p = <2e-16) were associated with faster publication (release of data). Presentation or conference materials were not associated with more timely dissemination in comparison to journal articles (p = 0.09). Studies that sampled blood donors/residual sera (p = 0.15) or that looked at multiple populations in one study (p = 0.26) did not differ in timeliness when compared to studies that investigated household or community samples; however, studies sampling healthcare workers (HR 0.81 [0.70 - 0.94], p= 0.004) and other special populations (HR 0.74 [0.64 - 0.86], p = 6e-05) took significantly longer to publish than studies that sampled the general population. There were no significant associations between timeliness and the WHO region a study was conducted in, when compared to AMRO. Further, there were no significant associations between timeliness and study representativeness or data validity, with the exception of the item evaluating if there was appropriate sampling or population adjustment. Adjusting for all other factors, non-probability sampling methods or not performing a population adjustment was associated with faster publication (HR 1.20 [1.03-1.41], p = 0.02).

## Discussion

Our analysis shows that many SARS-CoV-2 seroprevalence studies have not reported their findings in a timely fashion: with a median 154 days between sampling and reporting, there are considerable challenges in using these data for public health decision-making or impactful secondary analysis. Studies reported in preprints and peer-reviewed articles were much slower to be released compared to studies on other publication platforms, emphasizing delays introduced by the academic writing and publishing process that make seroprevalence studies less useful for public health decision-making and secondary analysis. However, we also show that it is possible to quickly release robust seroprevalence results; government or institution reports were more timely, and had better data validity and representativeness, compared to academic manuscripts. This suggests that there are opportunities to improve the timely reporting of strong seroprevalence studies and thereby improve their value for public health surveillance.

The slow reporting of SARS-CoV-2 seroprevalence studies overall emphasizes limitations in their relevance for public health action. The landscape of infection and immunity can change drastically in the median 154 days from the end of sampling to results release, particularly in an era of rapidly spreading SARS-CoV-2 variants and mass vaccination.^18^ Notably, some of the results from these studies are made available to public health agencies directly before being released publicly — for example, many studies of blood donors and residual sera in Canada. While this improves the ability of the agency in question to act on the data, the closed sharing of results hinders interpretation and action by other stakeholders. Firstly, public health agencies who the data has not been shared with (e.g., federal authorities, for studies done at a state/province level), which limits the coordination between levels of government that is crucial in a pandemic setting.^19^ Secondly, academic research groups, who have done secondary analysis and modeling that has generated key information during the pandemic.^20^ Finally, global synthesis and comparison initiatives: where this has been carried out for seroprevalence, these delays have caused limitations in the synthesis that can be done.^3^

We show that peer-reviewed manuscripts are released particularly slowly, with a median time-to-publication of about seven months. While many medical journals have expedited publication processes for COVID-19 research,^12^ our study demonstrates continued delay in the publication of seroprevalence findings. This raises the question of whether peer-reviewed journals are fit-for-purpose for reporting surveillance and seroprevalence findings. This is particularly true given that some journals may see routine updates on seroprevalence as not sufficiently novel to be published, potentially introducing publication bias.

The median preprint was reported close to four months faster than the median published article, but still nearly three months slower than the median government or institutional report. While preprints are enabling more rapid dissemination of information compared to publications, their median time to publication is still over three months, suggesting that the process of preparing an academic manuscript in the first instance introduces substantial delays. This suggests that pre-print platforms may themselves not be suitable for routine surveillance reporting.

In our analysis, studies of healthcare workers and other special populations took longer to publish compared to studies of the general population. This is in part because studies of the general population were typically regional or national studies conducted by government affiliated groups and disseminated via institutional or government reports,^10^ whereas studies of special populations were largely done by academic groups and released as research articles. However, these delays are problematic considering the importance of seroprevalence to inform best practices in high-risk settings, such as hospitals. There is a clear need for rapid release of the findings from these studies to enable their use for public health.

Interestingly, our analysis shows that it is possible for a seroprevalence study to be both timely and robust. Institutional reports, which were published rapidly, had a greater proportion of low RoB studies compared to preprints and published articles. Moreover, there were no differences in the timeliness of low, high, and moderate RoB studies on univariate analysis, suggesting that faster publication of valid study results is possible. However, studies with limited information to evaluate bias (unclear RoB) were published significantly faster. Many of these studies were published via news and media reports, which may account for the lack of data needed to evaluate bias. News and media outlets should endeavor to link to extended reports provided by investigators, even if not peer-reviewed or pre-printed on a formal platform, in order to maximize dissemination of crucial study context.

Collectively, these findings point back to a fundamental divide between the way in which research studies are ordinarily done and performing effective public health surveillance. Surveillance systems generate information for action, which may not be novel; in contrast, research generates information for knowledge, and is conventionally communicated via the peer-reviewed literature. The academic literature — whether peer reviewed or preprint — may not be fit-for-purpose as a way of communicating surveillance information. However, because many public health agencies have limited resources and serosurveillance expertise, they have often had to rely on results from intermittent studies conducted by academics — which are often delayed and not systematic. This approach is problematic, and accelerating peer-review processes may be unlikely to solve issues of timeliness.

Continuous serosurveillance, where governments perform routine serological sampling with rapid reporting, would address many of the challenges that the present study identifies. As an example, the REACT 2 programme involves repeated serological testing with an established analytical framework, thereby allowing for sampling, analysis, reporting, and data sharing.^11,21^ Continuous serosurveillance provides standardized, up-to-date data and avoids the need to extrapolate data over major gaps between sampling periods. To achieve this, public health agencies would need to be resourced to perform unique, ongoing, and systematic serosurveillance — either independently or in partnership with academic groups. However, these resources are often lacking, meaning that there are comparatively few examples of mature, well-funded, and ongoing serosurveillance systems run by public health agencies.

Recognizing that different public health systems and academic groups will conduct serosurveillance in different ways, there is a clear need for a centralized data system for serosurveillance data. This repository could serve as a modern version of the MMWR repository, which was initially created as a place to report surveillance data to enable public health action,^22^ and which we can improve on with modern technologies as GISAID has for genomic surveillance. Such an initiative could encourage standardization to a protocol such as the UNITY Study Protocols (WHO), with common data elements based on the ROSES Guidelines for reporting seroprevalence studies and serosurvey evidence synthesis efforts like SeroTracker’s.^1,23,24^ This would enable robust seroprevalence estimates to be rapidly deposited into a data repository, allowing expedited dissemination of data for immediate use for public health surveillance, secondary analysis, and synthesis. Rapid and flexible approaches to peer-review, such as crowdsourcing, could be built into such a repository to validate submitted data.^25^ Other efforts to generate surveillance data that have largely been done as research studies, such as point prevalence and wastewater studies could also benefit from such an approach.

Our analysis had several strengths. We aimed to identify all seroprevalence studies publicly reported in 2020 and 2021, providing complete coverage of seroprevalence studies in the first two years of COVID-19 pandemic. Our inclusion of studies across all geographic regions, publication venues, populations studied, and study designs enables comprehensive analysis. Moreover, the multivariate Cox regression conducted here enables us to isolate the association between timeliness and the covariates of interest, including study characteristics, measures of data quality, and measures of representativeness.

Some limitations of our approach should also be kept in mind. First, we were not able to determine the duration of key steps in the reporting process for each article — for example, to analyze samples, analyze data, prepare the report, peer review, and copyediting and typesetting. Greater granularity here would help inform tailored suggestions to expedite reporting. Second, we included the first public report of results for each seroprevalence study in our database. This avoids double-counting, but does not capture subsequent publications of that study in other venues. Our analysis showing that time to publication is similar for published articles regardless of whether they were first pre-printed suggests this had a limited effect. Third, some news and conference articles did not report an end date for sampling and had to be excluded. We also had to approximate sampling end date for studies reporting imprecise sampling dates (e.g., “between March and April”). Finally, studies that were reported directly to public health agencies and never publicly released could not be identified or included in this analysis.

Overall, our findings indicate that COVID-19 seroprevalence studies have often released results slowly through venues more suitable for research studies, limiting their utility as surveillance tools. It is crucial to prioritize the principles of surveillance in designing seroprevalence investigations. Well-resourced public health surveillance or close government-academic partnerships could help support continuous serosurveillance systems. At the same time, repositories for rapid and open dissemination of seroprevalence results can improve data comparability and enable secondary analysis. More timely, standardized, and robust reporting of seroprevalence results will increase their usefulness for surveillance, enabling more effective public health responses.

## Data Availability

The authors of this research article are contributors to SeroTracker, an interactive dashboard and data platform for SARS-CoV-2 seroprevalence. SeroTracker is a free, open-access tool that allows researchers and policymakers to visualize and analyze seroprevalence data. The data used in this research article was collected from the SeroTracker database which can be found and downloaded online (https://serotracker.com/en/Data).

## Abbreviations

RoB: Risk of Bias
JBI: Joanna Briggs Institute critical appraisal tool
FDA: U.S. Food and Drug Administration
EUA: Emergency Use Authorization
HR: Hazard Ratio
WHO: World Health Organization
COVID-19: Coronavirus Disease 2019

## Acknowledgements

We would like to acknowledge the SeroTracker Consortium, including the many research assistants who screened, reviewed and extracted seroprevalence study data as part of the SeroTracker living systematic review. We are especially grateful to Mercedes Yanes-Lane for her feedback during the conceptualization stage of this manuscript.

## Statement of Author’s Contributions

This is a secondary analysis of seroprevalence studies from the SeroTracker living systematic review database, maintained by the SeroTracker Consortium. CD and NI were co-first authors on this manuscript. Senior authors were DB, NB and RKA. **Claire Donnici:** Conceptualization, Methodology, Validation, Formal analysis, Investigation, Writing (Original Draft, Review & Editing), Visualization. **Natasha Ilincic:** Conceptualization, Methodology, Validation, Investigation, Writing (Original Draft, Review & Editing). **Christian Cao:** Conceptualization, Methodology, Investigation, Resources, Data Curation, Writing (Review & Editing). **Caseng Zhang:** Validation, Investigation, Writing (Review & Editing). **Gabriel Deveaux:** Validation, Investigation, Data Curation, Writing (Review & Editing), Graphical Abstract. **David A. Clifton:** Methodology, Writing (Review & Editing). **David Buckeridge:** Methodology, Writing (Review & Editing). **Niklas Bobrovitz:** Conceptualization, Methodology, Investigation, Writing (Original Draft, Review & Editing), Supervision, Funding acquisition. **Rahul K. Arora:** Conceptualization, Methodology, Resources, Writing (Original Draft, Review & Editing), Supervision, Project administration, Funding acquisition.

